# Post-acute sequelae of COVID-19 and adverse psychiatric outcomes: an etiology and risk systematic review protocol

**DOI:** 10.1101/2022.02.07.22270646

**Authors:** Andem Effiong

## Abstract

**Introduction:** The post-acute sequelae of COVID-19 (PASC) is a syndrome characterized by persistent COVID-19 symptoms or the onset of new symptoms following recovery from the initial or acute phase of the illness. Such symptoms often occur four or more weeks after being diagnosed with COVID-19. Although a lot of work has gone into understanding the long-term mental health effects of PASC, many questions related to the etiology and risk of this condition remain. Thus, this protocol is for a systematic review assessing the association between PASC and adverse psychiatric outcomes and whether people with PASC are at greater risk of developing an adverse psychiatric outcome than those without PASC.

**Methods and analysis:** Various medical databases (e.g., PubMed and EMBASE) will be searched for eligible articles using predefined search criteria. Gray literature will also be explored. Epidemiological observational studies and secondary analyses of randomized controlled trials that report a quantitative relationship between PASC and at least one adverse psychiatric outcome will be included. The Population, Exposure of interest, Comparator, and Outcome (PECO) framework will be used as a standardized framework for the inclusion criteria. The Joanna Briggs Institute (JBI) critical appraisal tools will be used to assess methodological quality and critically appraise the risk of bias in included studies. A random-effects meta-analysis will be conducted if possible. A formal narrative synthesis will be performed if a meta-analysis is impossible due to substantial heterogeneity across studies. The Grading of Recommendations Assessment, Development and Evaluation (GRADE) approach will be used to rate the cumulative certainty of the evidence for all outcomes.

**Ethics and Dissemination:** Ethical approval is not required for this study. The study results will be published in a peer-reviewed journal.

**Prospero registration number:** CRD42022308737

**Strengths and limitations of this study:**

▪ This study documents and addresses etiology, risk factors, and long-term symptoms of COVID-19 among people with post-acute sequelae of COVID-19 (PASC).
▪ It focuses on a key priority area for new evidence syntheses on the clinical management of COVID-19 and pandemic-related conditions.
▪ It will include evidence on non-hospitalized and hospitalized patients with a history of PASC.
▪ Substantial heterogeneity across studies may limit the ability to perform a meta-analysis.
▪ Findings will inform disease prevention, decision-making, healthcare policy, and clinical research.

## BACKGROUND

### Rationale

Coronavirus disease 2019 (COVID-19) is a contagious illness caused by the severe acute respiratory syndrome coronavirus 2 (SARS-CoV-2). Persistent and long-lasting (>4 weeks) symptoms following infection with acute COVID-19 have given rise to a syndrome known as post-acute sequelae of COVID-19 (PASC) or long COVID.^1, 2, 3^ Incidence and prevalence estimates for people with COVID-19 presenting with or reporting persistent psychiatric symptoms months following initial infection range from 0.8 - 49% ^1,2,3,4,5^ Among 44 759 people with no recorded history of psychiatric illness, the estimated overall probability of being diagnosed with new-onset psychiatric illness in the 90 days following a confirmed diagnosis of COVID-19 was 5.8% in a retrospective cohort study. ^6^ Similarly, clinical anxiety and depression, as well as other psychiatric sequelae, have been reported following diagnosis with COVID-19 in other studies.^6, 7, 8^ Although sex and age are considered to be sociodemographic risk factors for PASC, there is no consensus on other baseline clinical features that act as independent predictors of PASC.^9, 10^ The prevalence of PASC symptoms is higher in women compared with men.^10.^Among people aged 35-49 years, the prevalence of PASC is 26.8% compared with 26.1% and 18% among people aged 50-69 years and 70 years or older, respectively. ^10^

Persistent symptoms occur weeks and months after infection irrespective of initial disease severity (mild, moderate, severe, critical). ^11, 12^ Mendez et al. reported in their cross-sectional study that two months after discharge, neurocognitive impairment, psychiatric morbidity, and poor quality of life were markedly prevalent among 179 COVID-19 survivors who had been hospitalized.^12^ Nevertheless, Vannorsdall and Oh posit that current research on the post-acute phase following hospitalization has been conflicting due to the absence of a detailed, standardized neuropsychological evaluation of COVID-19 patients after hospitalization.^13^ In addition, they stated that literature on PASC and adverse mental health outcomes are mostly limited to studies that cannot establish causal relationships or lack generalizability (e.g., case reports, case series, and data obtained from cognitive screening instruments).^13^ Thus, more high-quality studies are warranted.^13^

In a study where the short-term and long-term sequelae of COVID-19 were systematically evaluated, PASC was categorized as short-term (1 month), intermediate-term (2-5 months), and long-term (≥ 6 months) following COVID-19 diagnosis.^14^ Clinical manifestations of PASC were classified into organ systems, i.e., cardiovascular, dermatologic, digestive, ear, nose, and throat; mental health, neurologic, and respiratory; constitutional symptoms; and functional mobility.^14^ The mechanisms leading to the post-acute and chronic neuropsychiatric manifestations of COVID-19 may be due to the direct effect of the viral infection and the indirect effect on mental health due to social isolation, post-traumatic stress, and job loss. Specifically, correlations have been observed between COVID-19 post-traumatic stress scores, general distress, and sleep disruption.^14, 15^ Despite those correlations, Khubchandani et al. stated that the causal pathways and etiology of adverse mental health outcomes in people who were infected with COVID-19 are multidimensional and complex.^16^

To clarify whether COVID-19 is a risk factor for psychiatric disorders and vice-versa, an electronic health record network cohort study of 69 million people consisting of 62 354 people with a COVID-19 diagnosis compared the rates of psychiatric sequelae of health in the initial four months of the pandemic (January – April 2020) and subsequently (after April 2020).^17.^ The study found that the rate of all diagnoses of psychiatric disorders and relapses was greater following a COVID-19 infection compared with after control health events (e.g., influenza infection, skin infection, other respiratory tract infections, and fracture).^17^ Similarly, a diagnosis of psychiatric disorder in the 12 months preceding the COVID-19 pandemic was associated with a 65% increased risk of COVID-19 (relative risk (RR) = 1.65, 95% confidence interval (CI): 1.59-1.71; p < 0.0001) compared with a matched cohort of people with specific physical risk factors for COVID-19 minus a psychiatric diagnosis.^17^ Whereas these associations were partly attributed to illness severity and pandemic-related contextual factors (e.g., social isolation, overwhelmed healthcare systems, and stigma), they do not adequately account for observed differences in psychiatric sequelae.^17^ Moreover, the inability to conclusively determine why there were between twofold and threefold increases in the risk of neurologic and psychiatric complications following a COVID-19 infection, in this and other studies, calls for further examination of the association between COVID-19 and risk factors for psychiatric morbidity.^17,18, 19^

With many long-term adverse mental health outcomes linked to COVID-19, effective interventions which optimize recovery and minimize relapse are needed. Such interventions may serve as appropriate tools to evaluate risk factors that may cause maladaptive psychiatric responses. ^20^ Furthermore, they may aid with the management of anxiety, fear, frustration, stigma, and paranoia by mitigating psychopathological symptoms and reducing contextual stress.^20^ Interventions that have been assessed in COVID-19 patients include virtual and physical psychotherapeutic approaches, e.g., cognitive behavioral therapy (CBT), emotional freedom techniques (EFT), and ultra-brief psychological interventions (UBPI); combined psychiatric and psychological interventions; technology and media; complementary and alternative therapies; self-care; spirituality and religion; and pharmacological.^21, 22^

Evidence on the effectiveness of these interventions is mixed and not thoroughly synthesized, with quality inadequately assessed in earlier studies and may vary depending on COVID-19 duration and severity. In a randomized controlled clinical trial of 51 people with COVID-19 consisting of an experimental group receiving progressive muscle relaxation (PMR) technology for 30 minutes each day for five consecutive days and a control group receiving only usual care and treatment, subjects in the experimental group reported lower depressive symptoms, lower anxiety levels, and better sleep quality compared with those in the control group. ^23^ Another randomized control trial of 30 hospitalized patients with COVID-19 assigned to an experimental or control group reported an improvement in all outcome measures among intervention group subjects compared to controls.^24^ In that study, a short four-session crisis intervention package tailored to cover COVID-19-specific guidance was delivered by clinical psychologists.^24^ Topics covered included tension reduction, relaxation, adjustment, responsibility skills enhancement, and promoting resilience.^24^ Outcomes measures in the study were derived from the Depression, Anxiety, and Stress Scale (DASS21), Symptom Checklist 25 (SCL-25), and the abbreviated version of the World Health Organization Quality of Life assessment (WHOQOL-BREF).^24^ Lack of cultural specificity, methodological issues, small sample sizes, lack of follow-up, unadjusted confounding factors, and brief time spans in both studies limit their generalizability.^23, 24^

During the COVID-19 pandemic, digital interventions to deliver health care have gained widespread acceptance.^25^ Remote care coordination and provision have been adopted to help reduce the risk of disease transmission.^25^ Mobile applications have also been used for contact tracing and information dissemination.^25^ Although an evidence synthesis of digital interventions to attenuate the adverse effects of the COVID-19 pandemic on public mental health highlighted their importance in mental disorder prevention and mental health promotion; it noted that evidence on their cost-effectiveness, process quality, and long-term outcomes is sparse. ^26^ Furthermore, the negative impact and risks of the COVID-19 pandemic are sometimes more significant in vulnerable and clinically extremely vulnerable populations (e.g., people over the age of 70, pediatric patients with cystic fibrosis, or people with developmental disabilities) who may be digitally disadvantaged.^26, 27, 28^

Presently, it is unclear what duration of PASC, etiologies and risk factors are most associated with the manifestation or persistence of adverse psychiatric outcomes (e.g., depression, anxiety, substance use disorder, post-traumatic stress disorder, psychosis, dementia, self-harm, suicide) compared with other health events. A prospective cohort study of patient-reported outcome measures (PROM) three months after initial COVID-19 symptom onset noted impairment with self-care and anxiety or depression as being present in 13% and 22% of its 78 subjects with at least one Charlson comorbidity at baseline compared to subjects without any Charlson comorbidities (4% and 9% respectively). Among subjects without any Charlson comorbidities, 70% reported an abnormal PROM, and 33% had at least one moderate issue in at least one EuroQol-5 Dimension (EQ-5D).^29^ In addition, questions remain about the long-term ((≥ 6 months) outcomes of COVID-19. ^30^

Although some studies indicate that most people who acquire COVID-19 are at risk of psychiatric sequelae and their symptoms tend to improve over time, others suggest that symptoms may worsen over time or point to a different disease trajectory.^30, 31^Research and any future recommendations about PASC and mental health should be guided by the best available evidence.

Few epidemiological studies have investigated the short and long-term impact of COVID-19 and PASC on mental health. Thus, this study will examine the causes of adverse psychiatric outcomes and risk factors in people with PASC. Furthermore, prior studies on this and related topics report internal validity and generalizability (external validity) limitations due to evidence derived solely from electronic health records, single network, or claims data. Because data on the psychiatric sequelae of PASC are conflicting and sparse, it is imperative to systematically summarize the evidence and combine the results of various scientific studies. This study aims to generate a new hypothesis on causality and provide a more precise estimate of the risk factors underlying PASC and adverse psychiatric outcomes.

An initial search of peer-reviewed and gray literature found no systematic reviews and meta-analyses on the topic. This protocol is for a systematic review that assesses the literature on PASC duration and risk factors that act as determinants (etiologies) of adverse psychiatric outcomes.

### Objectives

The primary objective of this systematic review is to determine whether people with PASC are at greater risk of developing an adverse psychiatric outcome (depression, anxiety, substance use disorder, post-traumatic stress disorder, psychosis, dementia, suicide) than those without PASC.

Secondary review questions include the following:

▪ Does the association between PASC and an adverse psychiatric outcome vary with age, sex, the severity of COVID-19 (mild, moderate, severe, critical), and duration of PASC (short-term (1 month), intermediate-term (2-5 months), and long-term (≥ 6 months) following COVID-19 diagnosis or hospital discharge)?
▪ Is PASC an independent risk factor for an adverse psychiatric outcome?

## METHODS

This protocol has been drafted following the Preferred Reporting Items for Systematic Reviews and Meta-Analyses (PRISMA) guidance for protocols (PRISMA-P).^32^ The systematic review will explicitly report any amendments and modifications made to this protocol.

### Eligibility criteria

#### Study design/characteristics

The review will include observational studies, namely, retrospective and prospective longitudinal cohort, case-control, cross-sectional, case series, and case reports. Secondary analyses of randomized controlled trials will also be included. Effect measures of risk factors, e.g., risk difference, relative risk, odds ratio, and hazard ratio central to the primary outcome, will be included. Risk factors predispose people with PASC to an adverse psychiatric outcome. Such risk factors are associated with an increased probability of people with PASC having a negative mental health outcome. Information on the relationship between risk factors and incidence of primary and secondary outcome measures will be included. Studies that do not report a quantitative relationship between PASC and at least one adverse psychiatric outcome will be excluded.

COVID-19 diagnosis must have been confirmed through clinical suspicion or with a positive nucleic acid amplification test (NAAT), e.g., reverse transcriptase-polymerase chain reaction (RT-PCR); antigen test; or serologic test (e.g., rapid serology test (RST) or enzyme-linked immunosorbent assay (ELISA)).^33^ Studies will be included if subjects were longitudinally observed since the initial diagnosis of COVID-19, i.e., during the acute phase or since the time of PASC onset (post-acute or chronic phase). A follow-up time of at least one month since the COVID-19 diagnosis is required. Primary and secondary outcomes will encompass etiology, risk factors, symptom and illness severity, duration of PASC, and adverse events.

#### Participants

Studies with adults as subjects (18 years or older) will be included. Pediatric and animal studies will not be included. There will be no sex, ethnicity, or race limitations. The search dates will range from December 2019 (date of first confirmed case of COVID-19) until March 2022 (the anticipated completion date of the review). COVID-19 filters will be used – if necessary – to limit search results to COVID-19 and PASC related articles.

#### Exposure

Primary measure

▪ Post-acute sequelae of COVID-19 (PASC), for this review defined as a continuing symptomatic illness or the emergence of new symptomatic illness in people with a confirmed history of COVID-19 after recovery from the acute phase of illness. PASC will be categorized as short-term (1 month), intermediate-term (2-5 months), and long-term (≥ 6 months) following COVID-19 diagnosis or hospital discharge.

Secondary measures

▪ Severity of COVID-19 (mild (including asymptomatic), moderate, severe, critical)

#### Comparators(controls)

Primary measure

▪ People with a confirmed history of COVID-19 without PASC

Secondary measures

▪ Severity of COVID-19 (mild (including asymptomatic), moderate, severe, critical)

#### Outcomes

Primary outcome variable

➢ Adverse psychiatric outcome
  ▪ Depression
  ▪ Anxiety
  ▪ Substance use disorder
  ▪ Post-traumatic stress disorder
  ▪ Psychosis

Secondary outcome variable

▪ Self-harm
▪ Suicide

### Information (evidence) sources and search strategies

Information, including titles and abstracts extracted from evidence sources, will be initially screened against the review questions. Information deemed eligible for inclusion will undergo more comprehensive screening. Once an article, study, or review is considered suitable for inclusion, it will be placed in the list of included studies. The steps above will be done for each information source, after which duplicates will be removed. The study selection process will be described in a PRISMA flow diagram and reported in the systematic review.

AE will develop the search strategy in consultation with a medical research librarian. The following databases and evidence sources will be searched: PubMed, Ovid MEDLINE, EMBASE, JBI EBP Database, CINAHL Plus, UpToDate, APA PsycInfo, Google Scholar, ProQuest Dissertations & Theses Global, Scopus, Web of Science, the University of Toronto COVID-19 Data & Statistical Sources, Centre for Addiction and Mental Health (CAMH) COVID-19 National Survey Dashboard reports, and COVID-END. Gray literature will also be considered where appropriate. Search strategies will be comprehensive and adapted for each information source. See Appendix 1 for a sample of the PubMed search strategy.

The Covidence (Covidence, Melbourne, Australia) or JBI SUMARI software will be used during the systematic review process for screening, appraisal of evidence sources, data extraction, synthesis, and study completion.

### Study selection

Information, including titles and abstracts, extracted from information sources will be initially screened by AE and a second reviewer against the research questions. Information deemed eligible for inclusion will undergo more comprehensive screening. Once an article, study, or review is considered suitable for inclusion, it will be placed in the list of included studies. The steps above will be done for each information source, after which duplicates will be removed. Disagreements on inclusion will be resolved through discussion or arbitration. The study selection process will be described in a PRISMA flow diagram and reported in the systematic review.

### Data extraction and management

Data will be extracted on primary and secondary outcome measures following the PRISMA guideline for systematic reviews.^35^ Outcome and effect size measures (e.g., adjusted and unadjusted odds, risk ratios, hazard ratios, standard errors), p-values, and associated 95% confidence intervals. RR for subgroups (e.g., age, sex, duration of PASC, COVID-19 severity) will be extracted if reported. The following data will also be extracted: authorship, publication year, journal name, study design, study location, sample size, baseline characteristics of subjects, demographics (age, sex, ethnicity, or race of subjects,’), study population characteristics (e.g., general population, prisoners, healthcare workers), the definition of PASC, duration of PASC, comorbidities, other risk factors, duration of follow-up, list of adjusted and unadjusted colliders (e.g., hospitalization, occupation, symptom recognition) and list of adjusted and unadjusted confounders (e.g., age, sex, nature of exposure, type of intervention), propensity methods.^34^ Two reviewers will conduct data extraction. Discrepancies in data extraction will be resolved through discussion or arbitration.

### Risk of bias in individual studies

The JBI critical appraisal checklist will be used to determine the methodological quality and critically appraise the risk of bias for included studies. Assessment will be done at the study and outcome level. Information related to a variable (exposure, outcome, covariate), misclassification, confounding, participant selection, reverse causation, missing data, study power, and generalizability will be appraised. Two reviewers will initially pilot the checklist to enhance consistency, mitigate potential issues with mechanistic scoring, and mitigate performance bias in the overall risk of bias assessment. Studies that do not adequately report on statistical analyses or address confounding, biases (selection, performance, detection, attrition), and other biases will be deemed lower quality studies, i.e., when they consistently have ‘no,’ ‘unclear, and ‘not applicable’ ratings’ across relevant items.

### Data synthesis

Summary treatment effects estimated as continuous outcomes will be converted to OR, RR, risk difference, and number needed-to-treat (NNT) with a 95% confidence interval (plus the baseline risk) for easier interpretation where possible. A random-effects meta-analysis will be conducted if possible. Statistical heterogeneity across studies will be explored using Higgins *I*^*2*^ and Cochran’s Q statistics. A Cochran’s Q test based on a χ2 statistic with a p <0.05 and greater than the degrees of freedom (df) will indicate heterogeneity. The *I*^*2*^ statistic will be interpreted as follows: 0-40% = minimal heterogeneity; 30-60% = moderate heterogeneity; 50-90% = substantial heterogeneity; 75-100% = considerable heterogeneity. If there is substantial heterogeneity, subgroup analysis (based on the duration of PASC or COVID-19 severity) will be conducted. Subgroup effect sizes (Cohen’s *d or* Hedges *g*) and correlations will be assessed and compared with unadjusted values to interpret for meaningful effects. Observed effects will be considered robust if the effect estimates of the primary outcome remain consistent or there are no large differences in the magnitude of effect across subgroups. Subgroup analyses will not be performed if there is minimal or moderate heterogeneity. A formal narrative synthesis will be performed if meta-analysis is not possible. The reasons for not pooling data (e.g., high statistical, methodological, and clinical heterogeneity) will be reported in the review. A methodological quality-based sensitivity analysis presented as a summary table will be used to assess the robustness of the findings. Authors of included studies with missing information will be contacted for clarification. The Grading of Recommendations, Assessment, Development, and Evaluations (GRADE) approach will be used to rate the overall certainty of the evidence obtained from the study.

### Patient and public involvement

Input on the review questions and outcomes was informally sought from patients and people who had been previously diagnosed with COVID-19 and PASC.

## Data Availability

All data produced in the present study are available upon reasonable request to the authors.

## Ethics and dissemination

Ethical approval is not required for this study. Study findings will be disseminated via preprints, peer-reviewed publications, conference abstracts, posters, plain language summaries, presentations, and infographics.

## Ethics statements

### Patient consent for publication

Not applicable.

### Contributors

AE conceived, designed, and drafted the study protocol.

### Funding

This work was supported by the Canadian Institute of Health Research (Grant number: NFRFR-2019-00012). The funding body will not have any role in the systematic review (and meta-analysis) process.

### Competing interests

None declared.

### Provenance and peer review

Not commissioned; externally peer-reviewed.

## Notes

### Competing Interest Statement

The authors have declared no competing interest.

### Author Declarations

This study involves only openly available human data obtained from PubMed and other medical databases.

### Summary of Updates

Background, Rationale, and Reference sections have been revised. The following updates were made: (1) Included the World Health Organization (WHO) and the National Institute for Health and CARE Excellence (NICE) definitions of PASC. See: Lines 51; 332-339. (2) The purpose of the study was revised for clarity. See: Lines 114-171.

