## Supplemental Table Appendix I for "Post-acute sequelae of COVID-19 and adverse psychiatric outcomes: an etiology and risk systematic review protocol"

**Supplementary material Appendix 1 PubMed search strategy (February 07, 2021)**

| #1 | (("SARS-CoV-2"[MeSH Terms] OR "COVID-19"[MeSH Terms] OR "COVID-19 breakthrough infections"[Supplementary Concept] OR "COVID-19 vaccine booster shot"[Supplementary Concept] OR "post-acute COVID-19 syndrome"[Supplementary Concept] OR "COVID-19 stress syndrome"[Supplementary Concept] OR "COVID-19 post-intensive care syndrome"[Supplementary Concept] OR "coronavirus disease 2019"[Text Word] OR "SARS-CoV-2"[Text Word]) AND "COVID-19"[Text Word]) OR "Risk Factors"[MeSH Terms] |
| --- | --- |
| #2 | "Mental Disorders"[MeSH Terms] OR "Diagnostic and Statistical Manual of Mental Disorders"[MeSH Terms] OR "Substance-Related Disorders"[MeSH Terms] OR "Neurocognitive Disorders"[MeSH Terms] OR "Patient Health Questionnaire"[MeSH Terms] OR "depress*"[Text Word] OR "anxi*"[Text Word] OR "substance use disorder"[Text Word] OR "posttraumatic stress disorder"[Text Word] OR "psycho*"[Text Word] OR "self-harm"[Text Word] OR "suicide"[Text Word] |
| #3 | "Causality"[MeSH Terms] OR "etiology"[MeSH Subheading] OR "Causality"[Text Word] OR "etiology"[Text Word] |
| #4 | #1 AND #2 AND #3 |
